# Diabetes and neuroaxonal damage in Parkinson’s disease

**DOI:** 10.1101/2022.04.19.22274027

**Authors:** Nirosen Vijiaratnam, Michael Lawton, Raquel Real, Amanda J Heslegrave, Tong Guo, Dilan Athauda, Sonia Gandhi, Christine Girges, Yoav Ben-Shlomo, Henrik Zetterberg, Donald G Grosset, Huw R Morris, Thomas Foltynie, PRoBaND clinical consortium

## Abstract

Patient’s with Parkinson’s disease (PD) with coexistent type 2 diabetes (T2DM) can manifest with more severe motor and cognitive phenotypes. The precise reasons for this remain unclear though underlying pathophysiological differences are increasingly implicated. More severe neuroaxonal injury in PD patients with T2DM was recently proposed as a potential mechanism for this. We explored this in the tracking Parkinson’s study and confirmed these findings. Cases with T2DM had higher serum neurofilament light levels and this remained significant after correction for age and vascular risk factor burden. Disentangling the underlying mechanism for more rapid axonal damage will be important in the development of disease modifying therapies for PD.

## Main text

We read with interest Uyar and colleagues’ recent report on the association between diabetes, non-diabetic elevated glycated hemoglobin levels (HbA1c) and neuroaxonal damage in PD patients from the MARK-PD study. (1) The authors confirmed previously established findings of an inverse association between diabetes and cognitive and motor status. The authors also demonstrated higher serum neurofilament light (NfL) levels (a marker of neuroaxonal damage) (2) in PD patients with prevalent type 2 diabetes and in PD patients with non-diabetic elevated HbA1c levels. These associations persisted after adjustment for age, body mass index (BMI) and vascular risk factors (prevalent arterial hypertension, hypercholesterolemia, and history of stroke). We recently noted similar motor and cognitive associations in PD patients with diabetes (3) in the Tracking Parkinson’s study though noted only a non-significant trend towards an association in the overall PD cohort between NfL levels and more severe motor and cognitive status at baseline (4) which may reflect the shorter disease duration in the Tracking Parkinson’s cohort, compared with the MARK-PD cohort.

Considering the authors’ novel findings of an association between diabetes and neuroaxonal damage, we explored the relationship between serum NfL and diabetes in our previously defined subgroup of the Tracking Parkinson’s study.(4) Analysis was performed using Stata V.17.0 (Stata, RRID:SCR_012763) and differences were compared using Kruskal-Wallis tests for continuous data and χ2 tests for categorical data while the association between NfL and diabetes was further explored using univariate and multivariate (age, BMI and vascular risk factors) linear regression analysis.

Of the 280 patients studied, 29 suffered from prevalent type 2 diabetes. PD-DM patients were older (74.1 years ± SD7.7 vs 68.1 years ± 8.7, p<0.001), with higher BMIs (31.1 ± SD5.7 vs 27.1 ±SD4.4, p<0.001) while a higher proportion had coexistent vascular risk factors than PD patients without diabetes(p=0.032). Serum NfL levels were higher in PD-DM patients (39.5 ± SD 18.9 vs 29.6 ± SD 16.0, p<0.001). Using regression analysis, NfL levels were significantly associated with patients’ diabetic status (Coefficient 0.82, 95% CI: 0.45-1.19, p<0.0001) which persisted (Coefficient: 0.52, 95% CI: 0.18-0.86p=0.003) following adjustment for age, BMI and vascular risk factors (history of angina, myocardial infarction, stroke, hypertension and hypercholesterolemia).

Our findings affirm Uyar et al’s report of an association between PD-DM and more severe neuroaxonal damage. Furthermore, the data indicate that the more severe phenotype in PD-DM noted to date by several studies is likely to be mediated by additional factors other than vascular risk factor burden that tends to co-exist in these cases. T2DM and PD share several pathological processes encompassing neuroinflammation, lysosomal dysfunction, mitochondrial dysfunction and the development of central insulin resistance that leads to neurodegeneration. (5) This process is in part mediated by hyperglycemia as demonstrated by the MARK-PD study and its downstream impact on alpha synuclein aggregation. (6) It is also possible that some of the observed associations are explained by diabetic neuropathy, as other peripheral neuropathies are known to increase blood NfL concentrations. (7) Disentangling the mechanistic factors which contribute to this more rapidly progressive axonal damage is of critical importance in the development of disease modifying therapies for PD.

## Data Availability

All data produced in the present study are available upon reasonable request to the authors

## Cohort studies

Tracking Parkinson’s is primarily funded and supported by Parkinson’s UK. It is also supported by the National Institute for Health Research (NIHR) Dementias and Neurodegenerative Diseases Research Network (DeNDRoN). This research was supported by the National Institute for Health Research University College London Hospitals Biomedical Research Centre and Cambridge BRC. The UCL Movement Disorders Centre is supported by the Edmond J. Safra Philanthropic Foundation.

## Biomarker analysis

Work on the biomarkers of progression in Parkinson’s and related disorders is supported by Parkinson’s UK and the PSP Association.

This research was funded in whole or in part by Aligning Science Across Parkinson’s [Grant ID: ASAP-000478] through the Michael J. Fox Foundation for Parkinson’s Research (MJFF). For the purpose of open access, the author has applied a CC BY public copyright license to all Author Accepted Manuscripts arising from this submission.

## Author’s Roles

1. Research project: A. Conception, B. Organization, C. Execution
2. Manuscript Preparation: A. Writing of the first draft, B. Review and Critique

Nirosen Vijiaratnam, 1A, 1B, 1C, 2A, 2B

Michael Lawton, 1C, 2B

Raquel Real, 1C, 2B

Amanda J Heslegrave, 1C, 2B

Tong Guo, 1C, 2B

Dilan Athauda, 1C, 2B

Christine Girges, 1C, 2BYoav Ben-Shlomo, 1C, 2B

Henrik Zetterberg, 1C, 2B

Donald G Grosset, 1A, 1B, 1C, 2B

Huw R Morris, 1A, 1B, 1C, 2B

Thomas Foltynie, 1A, 1B, 1C, 2B

## Author disclosures

### Financial Disclosures of all authors (for the preceding 12 months)

NV has received unconditional educational grants from IPSEN and Biogen, travel grants from IPSEN, AbbVie and The International Parkinson’s Disease and Movement Disorders Society, speaker’s honorarium from AbbVie and STADA and served on advisory boards for Abbvie and Brittania outside of the submitted work.

ML No competing interest

RR No competing interest

AJH No competing interest

TG No competing interest

DA No competing interest

CG No competing interest

YB-S No competing interest

HZ has served at scientific advisory boards for Abbvie, Alector, Eisai, Denali, Roche Diagnostics, Wave, Samumed, Siemens Healthineers, Pinteon Therapeutics, Nervgen, AZTherapies and CogRx, has given lectures in symposia sponsored by Cellectricon, Fujirebio, Alzecure and Biogen, and is a co-founder of Brain Biomarker Solutions in Gothenburg AB (BBS), which is a part of the GU Ventures Incubator Program (outside submitted work).

DG has received honoraria from BIAL Pharma, GE Healthcare, and Vectura plc, and consultancy fees from the Glasgow Memory Clinic.

HRM is employed by UCL. In the last 24 months he reports paid consultancy from Biogen, UCB, Abbvie, Denali, Biohaven, Lundbeck; lecture fees/honoraria from Biogen, UCB, C4X Discovery, GE-Healthcare, Wellcome Trust, Movement Disorders Society; Research Grants from ASAP, Parkinson’s UK, Cure Parkinson’s Trust, PSP Association, CBD Solutions, Drake Foundation, Medical Research Council. Dr Morris is a co-applicant on a patent application related to C9ORF72 - Method for diagnosing a neurodegenerative disease (PCT/GB2012/052140)

TF has received grants from National Institute of Health Research, Michael J Fox Foundation, John Black Charitable Foundation, Cure Parkinson’s Trust, Innovate UK, Van Andel Research Institute and Defeat MSA. He has served on Advisory Boards for Voyager Therapeutics, Handl therapeutics, Living Cell Technologies, Bial, Profie Pharma. He has received honoraria for talks sponsored by Bial, Profile Pharma, Boston Scientific.

## Notes

Financial Disclosure/Conflict of Interest: The authors declare that there are no conflicts of interest relevant to this work.

Funding Sources: HZ is a Wallenberg Scholar supported by grants from the Swedish Research Council (#2018-02532), the European Research Council (#681712 and #101053962), Swedish State Support for Clinical Research (#ALFGBG-71320), the Alzheimer Drug Discovery Foundation (ADDF), USA (#201809-2016862), the AD Strategic Fund and the Alzheimer’s Association (#ADSF-21-831376-C, #ADSF-21-831381-C and #ADSF-21-831377-C), the Olav Thon Foundation, the Erling-Persson Family Foundation, Stiftelsen för Gamla Tjänarinnor, Hjärnfonden, Sweden (#FO2019-0228), the European Union’s Horizon 2020 research and innovation programme under the Marie Skłodowska-Curie grant agreement No 860197 (MIRIADE), the European Union Joint Programme – Neurodegenerative Disease Research (JPND2021-00694), and the UK Dementia Research Institute at UCL (UKDRI-1003). DG has received grant funding from the Neurosciences Foundation, Michael’s Movers, and Parkinson’s UK.

### Competing Interest Statement

The authors have declared no competing interest.

### Author Declarations

The Tracking Parkinson's study has multicentre research ethics committee approvals

